# SARS-CoV-2 viral clearance and viral load kinetics in young children (1-6 years) compared to adults: Results of a longitudinal study in Germany

**DOI:** 10.1101/2022.08.09.22278540

**Authors:** Anna Sandoni, Angelika Schaffrath Rosario, Janine Michel, Tim Kuttig, Juliane Wurm, Stefan Damerow, Helena Iwanowski, Bianca Finkel, Livia Schrick, Udo Buchholz, Walter Haas, Gianni Varnaccia, Ulrike Kubisch, Susanne Jordan, Anja Schienkiewitz, Andreas Nitsche, Julika Loss

## Abstract

**Objective:** To investigate SARS-COV-2 viral clearance and viral load kinetics in the course of infection in children aged 1-6 years in comparison with adults.

**Methods:** Prospective cohort study of infected daycare children and staff and their close contacts in households from 11/2020-06/2021, comprising serial (self) sampling of upper respiratory tract specimen and testing for SARS-CoV-2 via PCR. Data on symptoms and exposure were used to determine the date of probable infection for each participant. We determined (a) viral clearance, and (b) viral load dynamics over time. Samples were taken from day 4-6 to day 16-18 after diagnosis of the index case in the respective daycare group (5 samples per participant).

**Results:** We included 40 children (1-6 years) and 67 adults (18-77 years) with SARS-CoV-2 infection. Samples were available at a mean of 4.3 points of time per participant. Among the participants, the 12-day study period fell in different periods within the individual course of infection, ranging from day 5-17 to day 15-26 after assumed infection.

Children reached viral clearance at a median of 20 days after assumed infection (95% CI 17-21 days, Kaplan Meier Analysis), adults at 23 days (95% CI 20-25 days, difference not significant). In both children and adults, viral load decreased over time with trajectories of the mean viral load not being statistically different between groups. Only small proportions of those tested positive had a viral load of >1 million copies/ml, which is considered the threshold for infectivity. Kaplan-Meier calculations show that from day 15 (95% CI 13-15), 50% of all participants that had a viral load no longer infectious or were negative.

**Conclusion:** Children aged 1-6 and adults infected with SARS-CoV-2 (wild type and Alpha variant) did not differ significantly in terms of viral load kinetics and time needed to clear the virus. Therefore, containment measures are important also in the daycare settings as long as the pandemic continues.

## INTRODUCTION

The role of daycare children in the spread of SARS-CoV-2 has been discussed controversially since the beginning of the COVID-19 pandemic in March 2020. Daycare centers promote the physical and psycho-social development, health and well-being of young children, and in many European countries reach a high proportion of young children from all social groups. In Germany, for example, 35% of 0- to 2-year-olds and 93% of 3- to 6-year-olds are enrolled in a daycare program (1). Pandemic-related closures or (repeated) quarantine and isolation periods of children are likely to have detrimental effects on psychosocial well-being, physical activity and body weight (2-5). However, daycare programs mainly serve children in an age group that is (for the time being, June 2022) not eligible for vaccination against SARS-CoV-2 in Germany. In addition, measures that help contain the transmission of SARS-CoV-2, such as wearing masks and physical distancing, are difficult to implement among toddlers and preschoolers. Therefore, understanding the transmissibility and kinetics of SARS-CoV-2 among daycare children is critical for the development of adequate mitigation policies, such as recommended time for isolation, testing strategies, and hygiene concepts contributes to understanding the role of daycare children in the spread of SARS-CoV-2 also in comparison to school children and adults.

Two indicators that help assess the infectivity of an individual and the duration of infectiousness are viral clearance (VC) and viral load (VL). VC is defined as the state when the virus is eliminated from the respiratory tract, as can be seen in negative PCR tests after a (series of) positive PCR test(s). VL refers to the amount of viral RNA detected in specimen from the respiratory tract. As we have learned in the course of the pandemic with regard to SARS-CoV-2, a positive PCR result does not equate to infectivity of the person tested. Rather, a threshold of SARS-CoV-2 RNA copies/ml has been determined above which it can be assumed that the amount of viral material is sufficient for transmission. Therefore, the measurement of viral load is helpful to better interpret PCR test results. Studies from the early period of the COVID-19 pandemic from Germany and the USA did not find significant differences between the VL of adults as compared to children aged 1-5 or 1-6 years, respectively (6, 7).

As for viral clearance, a comprehensive systematic review and meta-analysis, including studies published until 6/2020, yields a mean period of 17 days with a maximum of 83 days needed for COVID-19 patients of all age groups to reach VC from SARS-CoV-2 in the upper respiratory tract (8).

Most studies on VC and VL are based on data of hospitalized individuals or individuals seeking medical attendance (9-12). Mild or asymptomatic cases are probably underrepresented in these studies. This may especially be true for young children, as COVID-19 in childhood often does not present with any (severe) symptoms. We see a dearth of studies that (a) investigate the infectivity of SARS-CoV-2 positive individuals over time in a non-clinical community setting, (b) that correlate the PCR test results in the individual course of infection, and (c) explicitly focus on the age group of daycare children. Analyses have confirmed transmissions from infected children to close contacts within the daycare setting, and also to household members (13), highlighting the importance of investigating the role of daycare children in the infection process in more detail.

Therefore, we will examine the following research questions:

- How long does it take for children aged 1-6 to clear SARS-CoV-2 from their upper respiratory tract (VC)? Is there a difference to adults?
- How does the viral load (VL) of SARS-CoV-2 develop over time in children aged 1-6, compared to adults?

## MATERIALS AND METHODS

### Study design/Subjects

We analyzed data from the COALA study (14), which is an outbreak-related examination study in daycare centers, carried out from October 2020 to June 2021 in the “second” and “third wave”of then pandemic in Germany. At this time SARS-CoV-2 wildtype, followed by the alpha variant of concern (VOC), were predominant. COALA has a prospective longitudinal, case-related study design. Daycare center groups with one or more SARS-CoV-2 positive cases (child or staff) were enrolled in the study. 30 daycare groups from 20 different communities across Germany were included. SARS-CoV-2 cases as well as their close contacts within their respective daycare group and household were visited at home by a trained study team four to six days after the SARS-CoV-2 index cases’ test date. Further details of the study design and methods are described elsewhere (14).

A total of 1047 individuals was included in the COALA study (n=447 children and n=600 adults), of which only positively tested individuals, were included in the current analyses. Amongst the children, n=343 were in the age group of daycare children (1-6 years of age; children of older ages were siblings and not included here).

### Sampling

During the home visits, mouth-nose swabs (MNS) and saliva samples were taken. For the following 12 days after the home visit, participants were instructed to take these samples from their upper respiratory tract and from their children themselves every three days (self-sampling) and to send them to the Robert Koch Institute (RKI) by postal shipment (Fig. 1).

**Figure 1:**
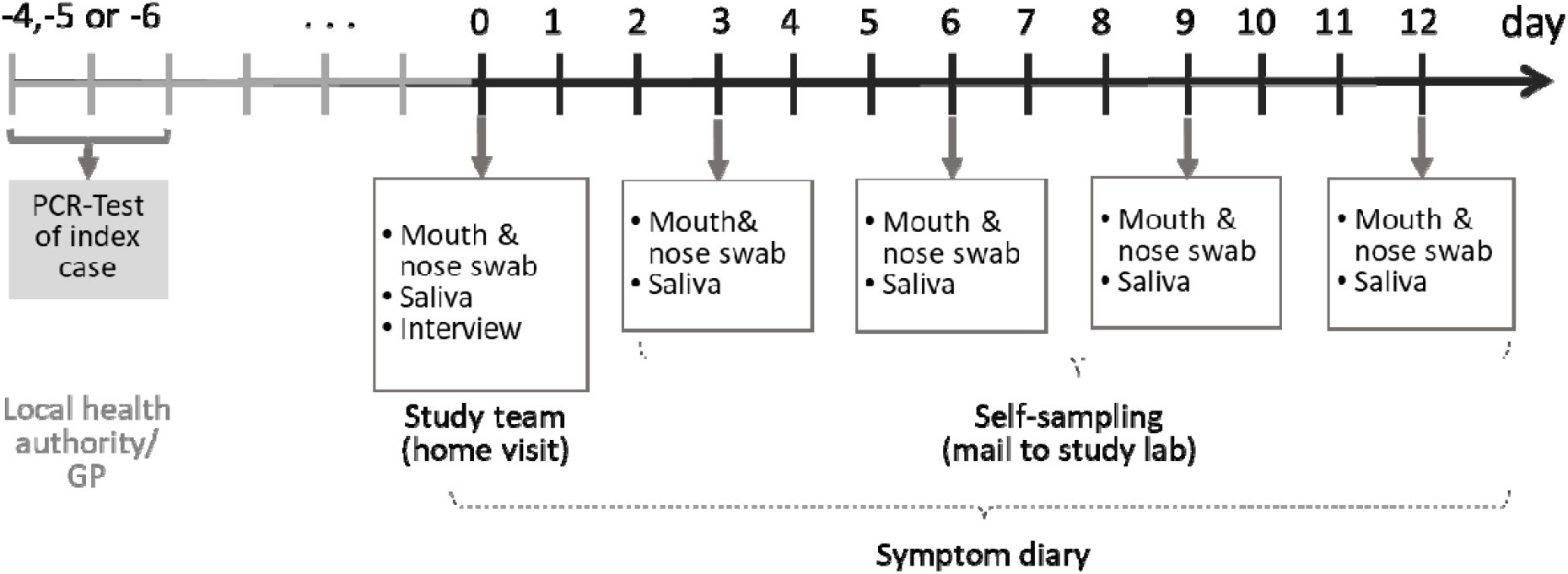
Timeline sampling scheme for bio samples taken from the participants of the COALA study (SARS-CoV-2 index cases, secondary cases and close contacts of SARS-CoV2 cases in the respective daycare center group and households). Participants were enrolled 4-6 days after the index case got tested. GP = general practitioner

**Figure 2:**
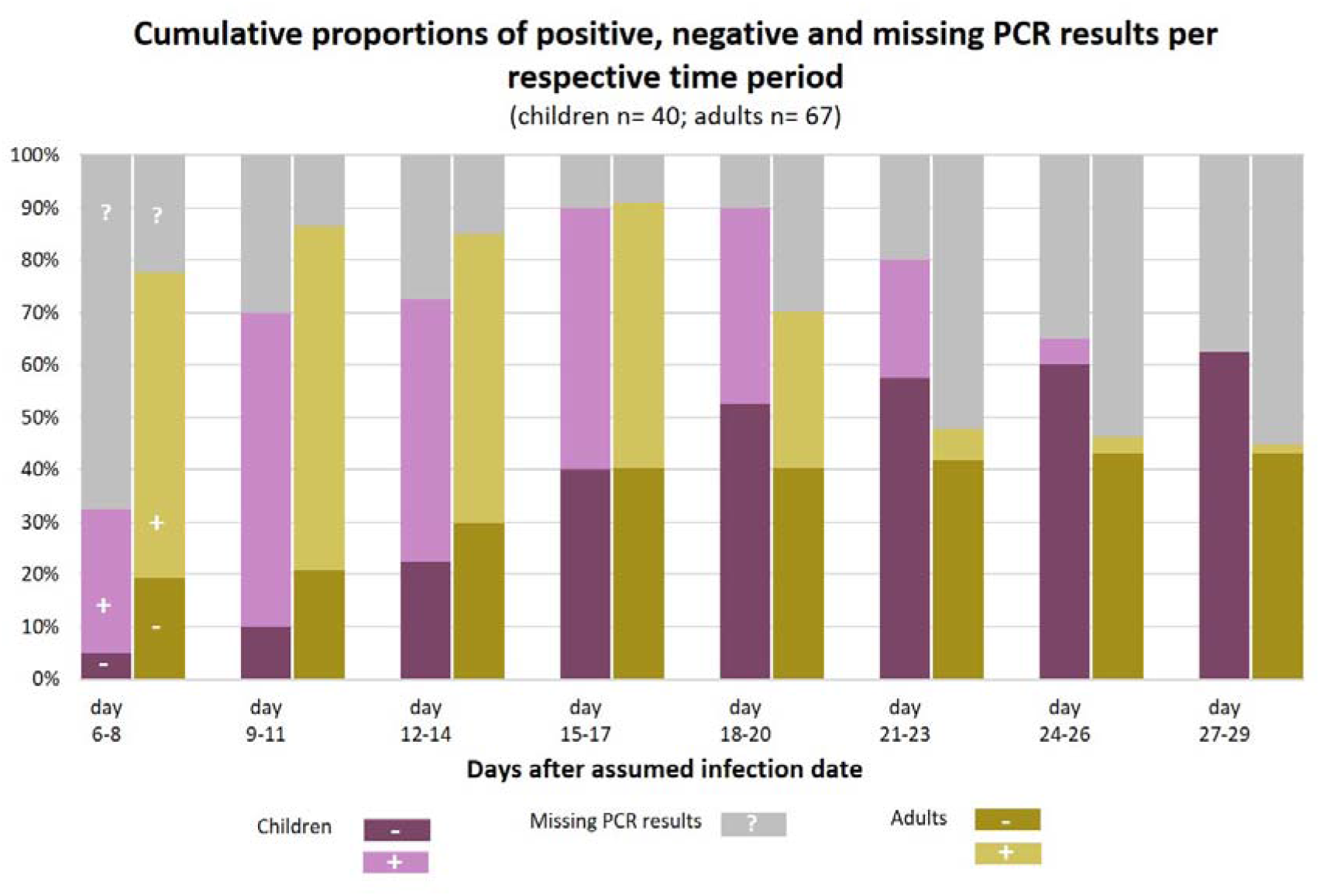
Cumulative proportion of children and adults with a negative (purple) PCR test on SARS-CoV-2, compared to the proportion of children and adults who were still tested positive (pink) at different points of time after the assumed infection. PCR test results were not available from all study participants at all points of time; the proportion of children and adults for whom no information of a SARS-CoV-2 PCR was available at a given point of time is shown in grey color. Reasons for missing PCR tests were for example that participants were not yet included in the study, e.g. index cases had probably been infected more than 6-8 days ago when the study team arrived for the testings (enrolment started 4-6 days after test date of index case). Other cases finished their 12-day self-sampling period earlier than day 20, or 24, or 27 after their infection date. In addition, few participants skipped single sampling appointments.

**Figure 3:**
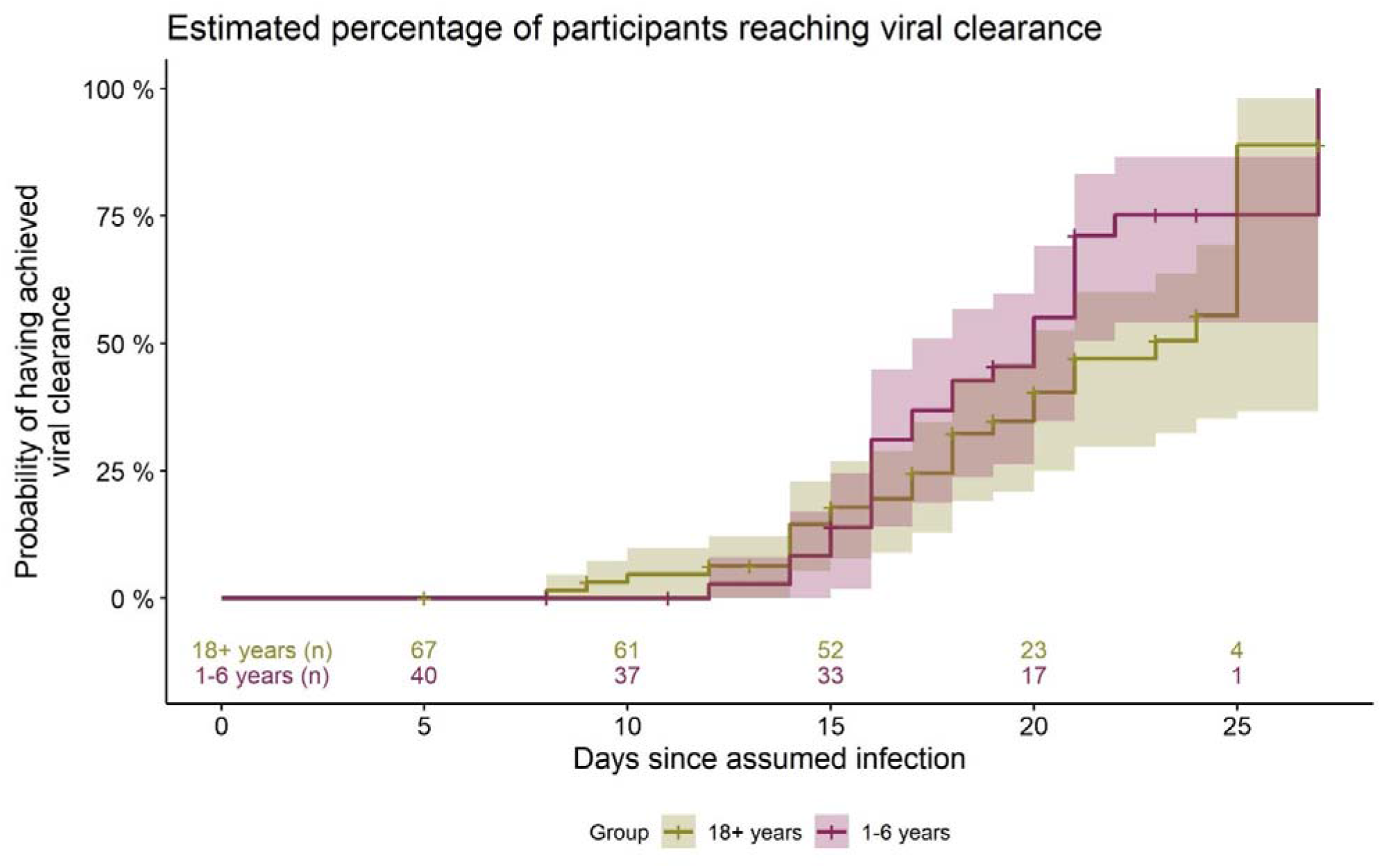
Estimated percentage (from Kaplan-Meier survival analysis) of daycare children and adults having accomplished viral clearance at the respective timepoints during the course of infection. Days counted since day of assumed infection (day zero). Group 18+ and 1-6 years: numbers of participants still positive immediately before the respective time point. Shaded area: 95% CI.

The close-meshed sampling and PCR testing over the 12 days study period made it possible to detect newly infected participants at an early stage. Moreover, it allowed to determine viral clearance and to quantify viral loads for each participant over the course of infection.

### Assessment of Symptoms

After the initial surveys during the home visits, household members were instructed to record clinical symptoms throughout the 12 days study period in a standardized symptom-diary, documenting symptoms e.g. fever ^≥^ 38°C, chills, cough, shortness of breath.

### Laboratory testing

All biological samples (MNS and saliva) were tested for SARS-CoV-2 RNA presence and quantity by real-time reverse transcription polymerase chain reaction (real-time RT-PCR) as described by Michel et al (16). Results were given in cycle threshold (CT) values (number of needed cycles surpassing the threshold for a positive test). For better comparison with other studies, CT values were converted to actual RNA copy numbers per milliliter. As a positive PCR result (values >=10^1 RNA copies/ml; ≙ CT 40 in our laboratory) does not equate infectivity, a threshold within the positive PCR range was defined above which an individual’s infectivity can be assumed. For SARS-CoV-2, this threshold was set at 10^6 RNA copies/ml (≙ CT 24.7 in our laboratory) (17). Log10 RNA copies/ml were calculated based on the cycle threshold (CT) of the E gene. Comparative analysis of E gene and delta CT of E gene and c-myc, either obtained in the multiplex or in a single plex PCR, showed no significant difference in exemplary sample courses, allowing the use of the E gene CTs directly for calculation of RNA quantities (exemplary courses shown in Suppl. fig 1).

### Determination of assumed infection date

In order to determine the starting point for calculating the period until viral clearance is reached, we decided against using the onset of symptoms (as young children are often asymptomatic or present only with very unspecific symptoms). Neither did it seem recommendable to use the date of the first positive PCR test, as the timepoint of testing among the cases differed substantially (e.g. sometimes asymptomatic primary cases were not detected before secondary cases who presented with symptoms were diagnosed). We therefore decided to determine the probable date of infection with SARS-CoV-2 for each participant, using date of testing, symptom diary, and questionnaires on exposure and symptom history. All sampling days were then expressed as days since assumed infection for each participant. For calculations, the index cases’ first positive test date by the health department and a mean incubation period of SARS-CoV-2 of six days were taken as a basis and applied to index cases and secondary cases in daycare centers and households.

### Definition of VC

To determine viral clearance, we calculated the time from the assumed date of infection until the first negative PCR test, which was not followed by another positive PCR test in the self-sampling. If only one of the two samples received (MNS or saliva) was positive, the person was classified as positive at that time.

### Definition of VL

Viral load is defined as the number of SARS-CoV-2 RNA copies/mL in the PCR sample. If both the MNS and the saliva sample of a person had a positive result on the same day, we used the sample with the higher viral load for the analyses.

For the analysis of viral load, CT values measured in the PCR analysis were converted into RNA copies/ml by the following formula: 83.33333 * e^ ((CT value – 38.248)/–1.4) that was determined by serial dilution and measurement of pre-quantified target RNA. Then, the number of RNA copies/ml was transformed to the log scale (with base 10), resulting in a linear transformation of the original CT values, but with opposite sign.

### Statistical analysis

To examine the time to viral clearance, Kaplan-Meier survival time analysis was applied. The time from infection to the first negative test is equaled with “survival” time. A Kaplan-Meier analysis computes the probability of occurrence of a negative PCR-test at a certain point of time, taking into account the number of participants still in the study at this time. For some participants, a negative test was not yet recorded during the study period, i.e. they still had a positive test on the last sampling day. In these cases, the event of interest (PCR negativity) did not happen during this time. These participants were labeled as “censored observations”. For these censored observations, the only information is that the event (=first negative test) did not occur by the end of the study, which can be due to the following reasons: a) inclusion in the study early in the individual course of infection, e.g. when a secondary household case got infected during the last days of the study period, b) early study dropouts and c) prolonged viral shedding.

To test for significant differences between daycare children and adults, a log-rank test was applied, which is the standard test for comparison of two groups in Kaplan-Meier survival time analysis.

For viral load over time, mean, median, quartiles and distribution were calculated separately for children and adults. The difference in the mean course of viral load for children vs. adults is given as mean (Fig. 4) and median with 95%-confidence interval (Fig.5) for each time point. The difference was tested for significance in a linear mixed model which included VL as dependent variable, the individual participant as random effect and day since assumed infection (as a linear variable), age group (child/adult) and specimen type (mouth-nose swab or saliva) as independent variables. Descriptive calculations were performed using STATA 17.0 (Stata Corp, College Station, Texas, USA, 2021). SAS 9.4/TS1M7 (SAS Institute Inc., Cary, NC, USA, 2016) was used for survival analysis and mixed model calculations and R 4.1 (R Foundation for Statistical Computing, Vienna, Austria, 2022, www.R-project.org) was used for graphical representation.

**Figure 4:**
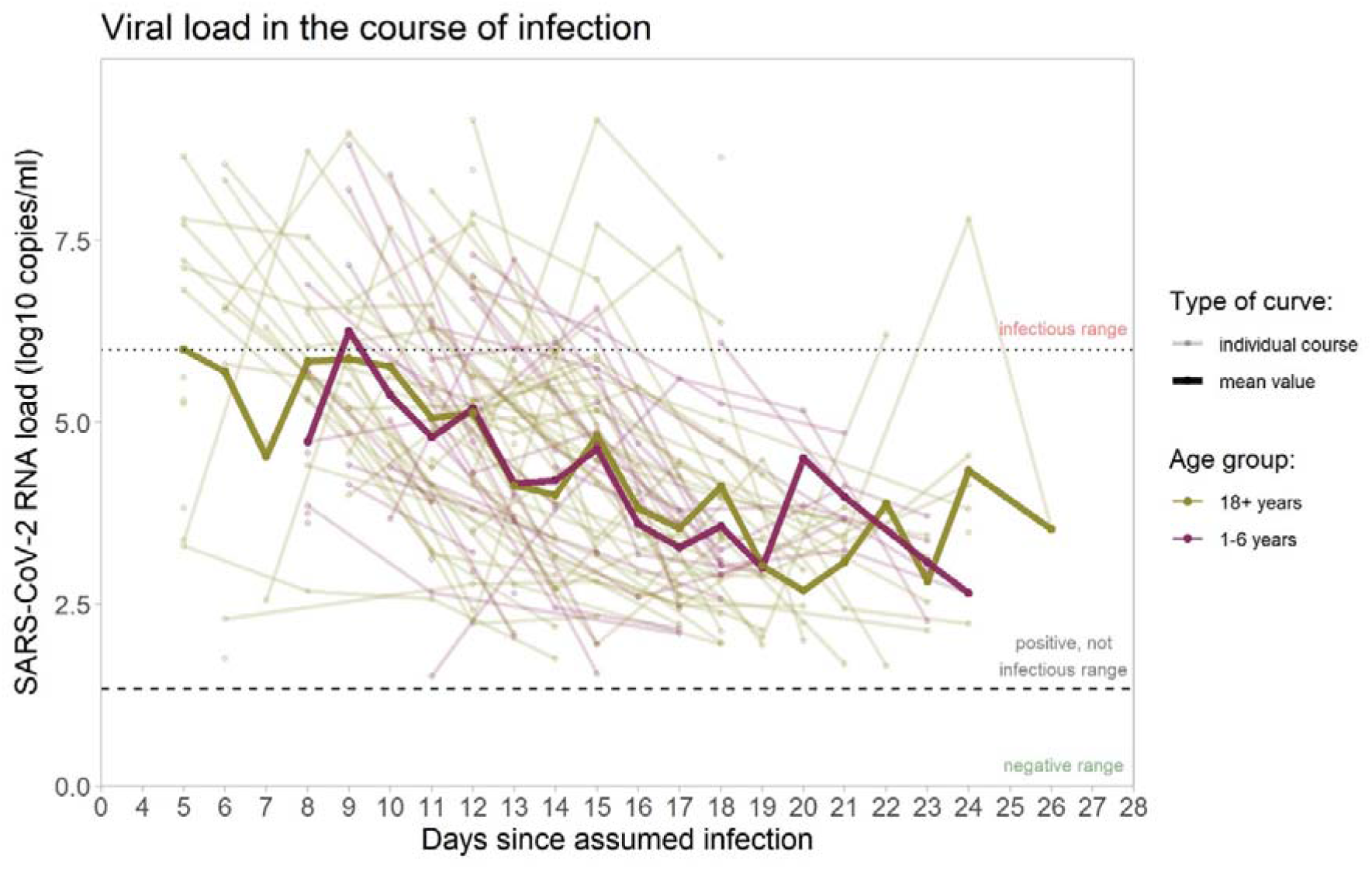
SARS-CoV-2 RNA copies/ml in the samples from upper respiratory tract. Viral load courses (individual courses and mean courses of children and adults), shown from the day of assumed infection (day zero) onwards. Dotted line marks 10^6 RNA copies/ml as threshold for infectivity (>=10^6 RNA copies/ml denotes infectious range); dashed line marks threshold for positivity with values >=10^1 RNA copies/ml considered as positive. Analysis restricted to samples considered positive (102 samples in 40 children, 214 samples in 67 adults) with increasingly fewer samples in the progression of the course of infection

**Figure 5:**
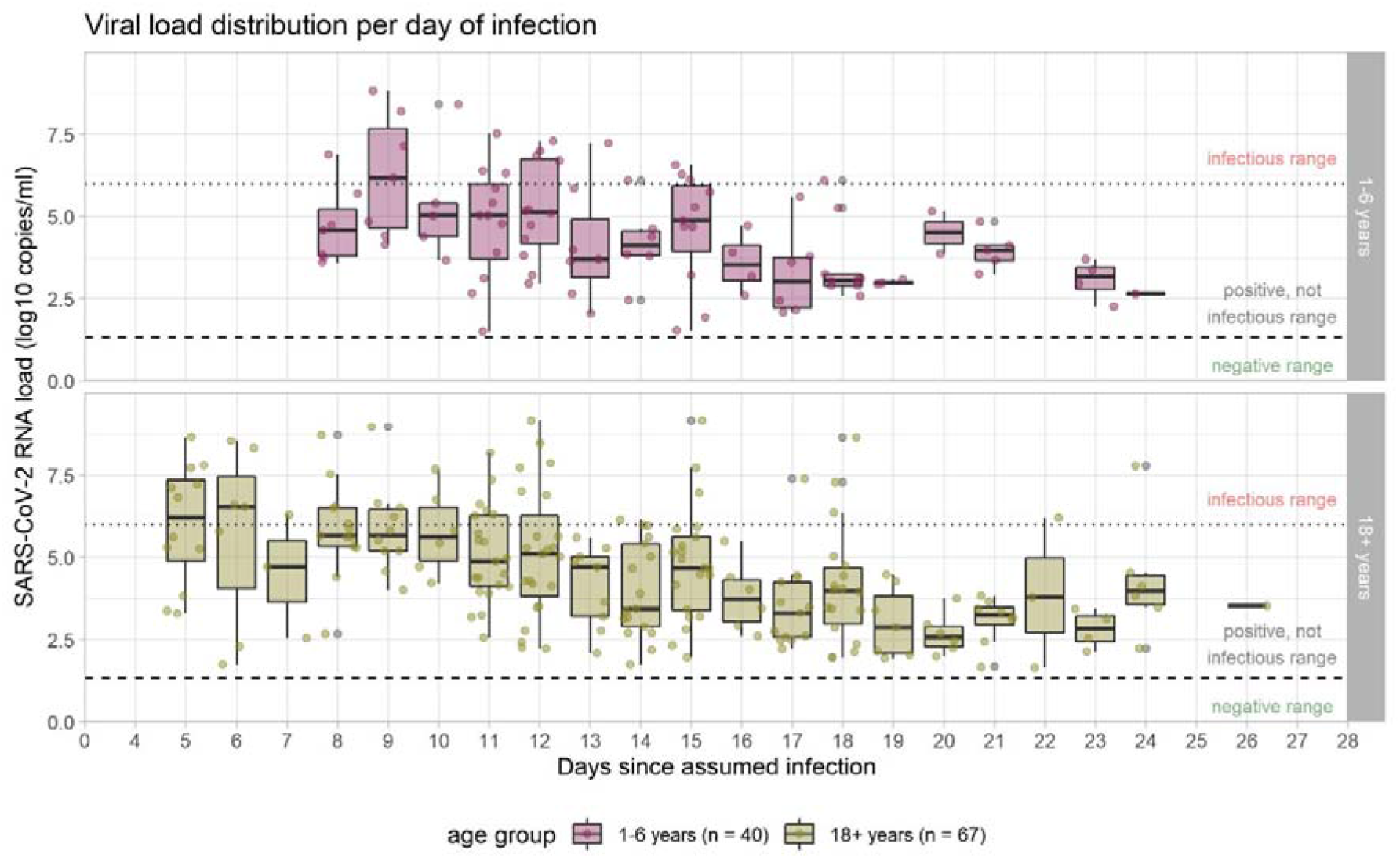
Viral load in the course of infection. Medians are calculated for those samples which were available on the given days, and are indicated by midlines; the top and bottom edges of boxes represent the interquartile range (IQR). Whiskers indicate the upper and lower values. Dotted line marks the threshold for infectivity 10^6 RNA copies/ml (≙ CT 24.7 in our laboratory); dashed line marks the threshold for positivity with values >=10^1 RNA copies/ml (≙ CT 40 in our laboratory) considered as positive. Analysis restricted to samples considered positive (104 samples in 40 children, 214 samples in 67 adults) with increasingly fewer samples in the progression of the course of infection.

## RESULTS

### Sample

Of all included individuals, 67 adults (18-77 years; female=56,7%) and 40 children (1-6 years; female=50%) were tested positive by PCR for SARS-CoV-2 in at least one specimen during the COALA study and were included into the current analysis.

### Viral clearance

26/40 children and 29/67 adults reached viral clearance during the study period, with the last sampling occurring on day 8 to 30 (children), and day 5 to 27 (adults), respectively, after their individual infection date. The remaining 14 children and 38 adults still had positive PCR results at the time of the last sampling in the study period. Therefore, no statement on VC can be made for these participants on an individual basis, but they can be included in the Kaplan-Meier analysis.

Figure 2 shows the cumulative proportion of children and adults who were found to have cleared SARS-CoV-2 at different points of time after infection.

According to Kaplan-Meier-survival time analysis, the estimated median time from infection to the first negative PCR test, i.e. time to viral clearance, is 20 days (95% CI 17-21 days) for the daycare children. 1/4 had their first negative test at or before an estimated 16 days after infection, 3/4 at or before day 22. The percentage of children having reached VC at day 21 was estimated as 71% (95% CI 55-85%).

In the adult cohort, viral clearance occurred at an estimated median of 23 days after infection (95% CI 20-25 days). 1/4 had their first negative test at or before an estimated 18 days since infection, 3/4 at or before day 25. The percentage of adults having reached VC was 47% by day 21 (95% CI 33-63%), 56% by day 24 (95% CI 40-72%) and 90% by day 25 (95% CI 63-99%) (Fig. 2).

The log-rank test for differences between children and adults has a p-value of p = 0.10 (χ² (1) = 2.65), showing that distributions between the groups do not differ significantly.

### Viral load

Changes of viral load over time, throughout the natural course of infection, vary substantially between individuals (Fig. 4).

The broad range of all viral load values of different individuals on certain days after infection is also reflected in the boxplot figure (Fig. 5). Regarding the raw mean values of viral load, there is a decreasing trend over time since infection, with no clear differences between children and adults. The mean viral load of both groups (adults and children) stays below the threshold of 10^6 RNA copies/ml, considered necessary for transmission, from day nine the course of infection. The same holds true for the median (Fig. 5), indicating that at any time point after day nine, less than half of the positive specimens (often, considerably less than half) were above the infectivity threshold. Single values above this threshold were observed until day 18 in children and until day 24 in adults.

Furthermore, the linear mixed model indicates an estimated difference in the trajectories of the mean viral load over time of -0.35 log10 RNA copies/ml (95% CI -0.79-0.10) for children vs. adults (p=0.12) which is not statistically significant.

Among the participants with a positive PCR test result, those whose viral load is considered high enough to allow virus transmission to others, are of special interest from a public health point of view. Table 1 details that only small proportions of those tested can be considered infectious, confirming graphic evidence from Figures 3 and 4. The proportion of individuals in whose samples more than 10^6 SARS Co-V-2 RNA copies/ml could be detected, peaked until days 11 and 12, and then decreased markedly (Tab. 1). Kaplan-Meier calculations show that from day 15 (95% CI 13-15), 50% and from day 17 (95% CI 15-20), 75% of all participants had a VL that can be classified as either negative or no longer infectious (graph not shown).

**Tab 1:**
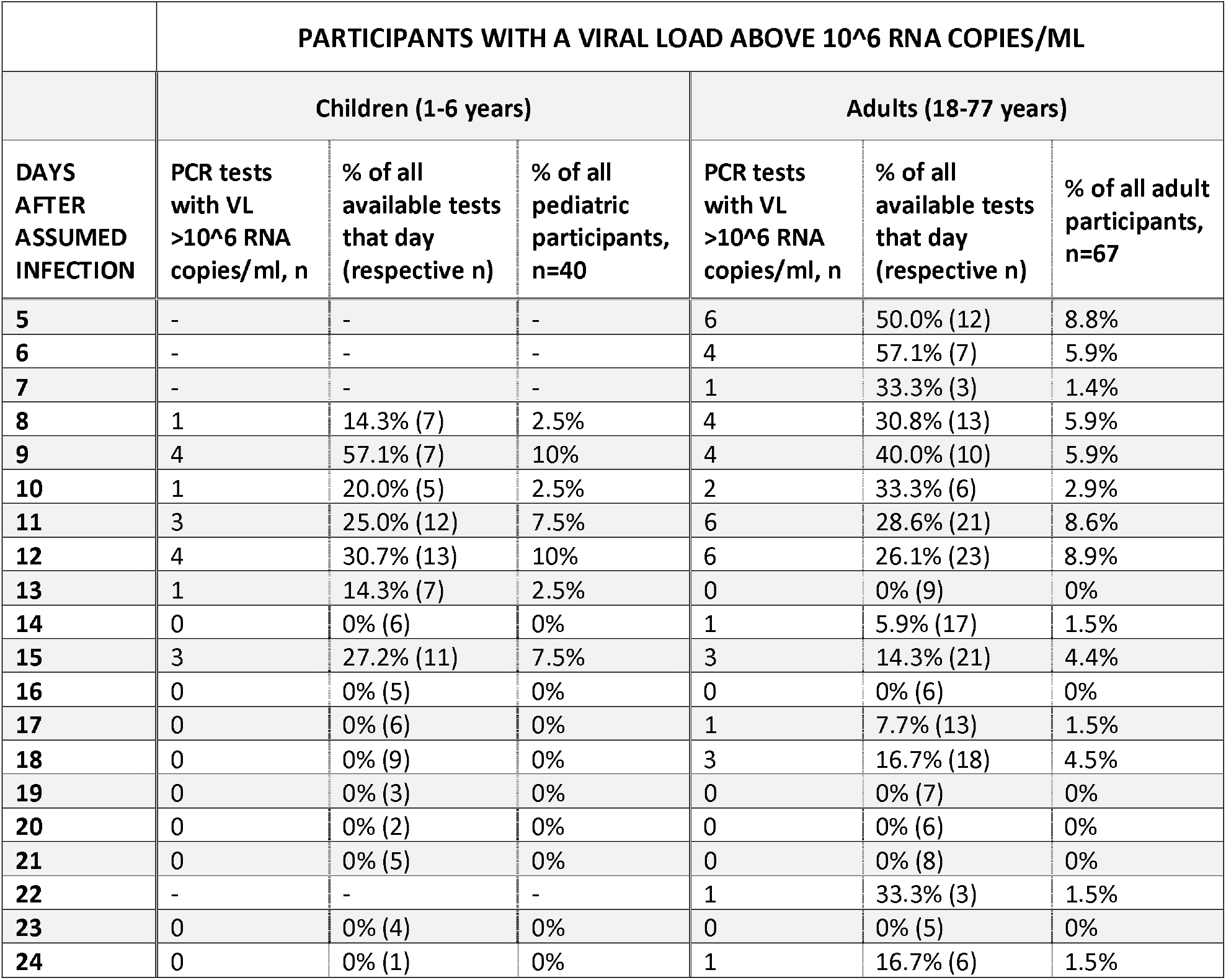
Number and proportion of children and adults showing a viral load in the infectious range, detailed for different days of the infection course. Samples are not provided by all participants on each day, because (a) the sampling scheme asked participants to send in swabs every third day only, and (b) the sampling scheme started on day 4-6 after testing date of the index case, therefore participants were included on different days of their individual course of infection, and the sampled over a period of 12 days. Dashes: no samples received.

Overall, more than half (n=63, 59%, 95% CI 49-68%) of the participants with positive SARS-CoV-2 test did not show a VL considered to be infectious at any day of the study period. Numerical differences between children and adults were not significant (χ²-test: p= 0.32).

## DISCUSSION

### Summary of main findings

Among daycare children (age 1-6 years), the median time span from assumed SARS-CoV-2 infection to the first negative PCR test was 20 days, which was not significantly different from the age group of adults (23 days). Presuming an incubation period of six days, this may roughly be translated into a median period of 14 days after onset of symptoms (if any) until the virus is cleared from the respiratory tract. Although SARS-CoV-2 may persist for weeks and viral loads throughout the individual course of infection are very heterogeneous, its quantity in the airways is mostly below the threshold set for an individual’s infectivity, as could be shown by tracking of viral load over time. 17 days after assumed infection (i.e. circa 11 days after symptom onset), 75% of participants were not infectious anymore, as they had either a negative PCR test or a positive test with a viral load of less than 1 million copies per milliliter. Viral load, in the mean, decreased gradually over time, and the viral load trajectory did not differ significantly between young children and adults.

Further findings from the COALA study, which focused on actual transmission risk and secondary attack rates (13) also underscore no significant differences between daycare children and adults: In the enrolled daycare centers, the transmission risk from pediatric primary cases did not differ significantly from that from adult primary cases (11% vs. 7.0% of close contacts got infected).

### Comparisons with other studies

Our study confirms the finding that SARS-CoV-2 is often present in the upper airways for weeks, while other viruses such as influenza are rarely shed for more than seven days (8, 18-20).

Prolonged periods of continued or inconsistent positive testing may be explained by the physiological half-life of respiratory epithelial cells (three months) and the ability of the PCR tests to detect non-viable viral fragments at very low concentrations (21, 22).

When comparing our results with those of other SARS-CoV-2 studies, it should be reflected that there are different methods to calculate the elimination period. VC, i.e. the state when a person has ‘cleared’ the virus from their body, is sometimes defined as the day of the last positive PCR test, in other studies as the day of the first negative PCR test. For calculating the period until VC is reached, studies have referred to the day of symptom onset, or the day of the first positive PCR test. Therefore, calculations of the period needed for an individual to eliminate the virus can only cautiously be compared between different studies.

Regarding VC we found three relevant studies: Gupta et al. (23) analysed data of 85 children and adolescents (aged 0-18) who were tested positive for SARS-CoV-2 and admitted to hospital during April and May 2020 in Rajasthan/India. The participants were tested by PCR every 3 days. The authors found that median time for children to clear the virus was 7 days, calculated from symptom onset, which is considerably shorter when compared to our results. Gupta et al. found that 99% of children reached viral clearance by day 15 after symptom onset; this may correspond to approx. 19-21 days after infection, a period after which, in our study, only approx. 50-75% of children were free of virus, according to the Kaplan Meyer analysis.

A German study among 208 participants carried out between January and May 2020 (24) showed results similar to ours. Here, the median time to reach VC was 12.5 days, and 75% of the participants were determined free of SARS-CoV-2 21.5 days after symptom onset. However, the study had not focused on young children, but included participants of all age groups, without reporting on differences between age groups.

A notably longer median time to clear the virus from one’s body was found in a study conducted from March-June 2020 in Washington/USA. Median time to reach VC (from first positive PCR test) was 22 days for children aged 0-5 years (n=24) (25). Overall, the elimination period in the different studies shows inconsistent duration of about 13 to 24 days for VC, and our result of 20 days fits well with the current state of research.

Three studies were found that assessed individual VL of SARS-CoV-2 over time: In the sample presented by Bahar et al., there was no meaningful difference between viral loads throughout the course of SARS-CoV-2 infection between children and adults (25), which is similar to our finding. Costa et al. investigated 256 infected children (1-18 years) and 928 adults (from June 2020-January 2021 in Spain) and found similar SARS-CoV-2 viral loads in the first days after symptom onset. However, PCR tests after three days after symptom onset showed a significantly lower VL in children as compared to adults (p=0.002) (26). Our results could not directly confirm this finding, but this may partly be due to the smaller sample size.

Jang et al. also looked into the progression of VL, and describe that from about day 10 post-infection, SARS-CoV-2 RNA can still be detected, but its amount remains predominantly below an infectious threshold (27). This finding is consistent with the viral load dynamics found in our study.

### Limitations/strengths

First, it is a limitation that the sample size of the COALA study is relatively small, and may not be representative for all children infected with SARS-CoV-2 in this age group. In addition, the participating adults were recruited from the daycare setting, which means they were mostly staff or parents of young children. On the other hand, this sample can provide insights into courses of SARS-CoV-2 kinetics outside the clinical setting. The participants were closely examined over a period of almost two weeks. Thus, compared to other studies, it was possible to analyze viral load dynamics much more precisely, and to identify SARS-CoV-2 cases that might not have been detected otherwise, for example due to a lack of symptoms.

Second, as our study design focused on the role of children, we included pediatric index cases preferentially to adult index-cases. As sampling started at the same point of time for primary and secondary cases, primary cases were more likely to be tested in later stage of infection. Therefore here were fewer censored observations in the group of children than in the adult group which may bias the statement regarding VC duration and VL.

Third, as the data collection for our study was set from 10/2020-6/2021, when the SARS-CoV-2 wild type and alpha VOC were dominant, it is unclear whether conclusions can be drawn about other variants.

### Implications for policy and practice

The observation that it usually takes several weeks for children aged 1-6 years to eliminate SARS-CoV-2 from one’s respiratory tract emphasizes that SARS-CoV-2 cases in daycare programs among enrolled children should be taken seriously, and isolation for infected children and quarantine of (non-immunized) close contacts need to be adhered to for an adequate span of time.

Our data also show, on the other hand, that persisting positive tests may not necessarily correspond to a child’s infectiousness. In our study the vast majority of positive SARS-CoV-2 PCR test results beyond day 15 after the probable infection yielded a viral load well under the threshold of 10^6 RNA copies/ml. Viral load is recognized as a strong determinant of transmission risk (28) and according to our data, there are probably only few days when a child’s dose of viral shedding is high enough to transmit the virus to others. These findings may add to other data which inform decisions on the length of isolation periods for children and adults. Other factors besides viral shedding (type of contact, respiratory volume, coughing, etc.) also play explicit roles in transmission, and viral shedding alone cannot be equated with transmission. In addition to epidemiological research findings on SARS-CoV-2 transmission in daycare-aged children, psychosocial and educational aspects should be considered for the selection of appropriate containment measures in the daycare setting.

## Conclusion

On the whole, our study does neither fuel fears that children may be significant drivers of the COVID-19 pandemic, nor confirm theories that they hardly play a role in transmission dynamics. The viral load kinetics and transmission risks of young children are similar to those of adults, which supports to maintain containment measures in the daycare setting in order to provide protection to children and staff members.

## Data Availability

All data produced in the present study are available upon reasonable request to the authors

## Ethics statement

The Ethics Committee of the Berlin Medical Association has reviewed the COALA study and approved the implementation of the study (Eth-39/20). The participation in the study is voluntary and all participants are informed about the objectives and contents of the study as well as data protection and give their written informed consent; children aged from 14 to 17 years give their own written consent in addition. Every participant is assigned to a sequential study number (ANR) in order to ensure pseudonymization of the study documents and material.

COALA is registered in the German Clinical Trail Register (DRKS-ID=DRKS00023501).

## Acknowledgements

We would like to thank all our colleagues at the RKI in the Department Epidemiology and Health Monitoring, the Department Infectious Disease Epidemiology, and the Centre for Biological Threats and Special Pathogens for their support. Special thanks to the colleagues of SurvAd, the Epidemiological Central Laboratory, and the COALA study team at the RKI. We would like to thank all participants for their support of the study. For their advice in the development of the study design we would like to thank all federal state health authorities, local health authorities, experts on day care centers, and colleagues at the German Youth Institute (DJI) who supported us designing the interviews.

## Author contributions

JL, ASch, UB, WH and SJ designed the study. JWu, HI, ASa, UK and BF were responsible for participant recruitment, data and specimen collection. AN, JM, LS performed laboratory testing and analysed lab results. ASR, JWu, SD, ASch, TK, ASa, and GV carried out the analysis and were responsible for the accuracy of the data analysis.

ASa, JL and ASR wrote the manuscript, which was then reviewed and approved by all other authors.

